# “COVID-19 in twins: What can we learn from them?”

**DOI:** 10.1101/2021.09.29.21263145

**Authors:** Mateus V de Castro, Monize V R Silva, Michel S Naslavsky, Keity S Santos, Jhosiene Y. Magawa, Edecio Cunha Neto, Maria Rita Passos-Bueno, Mayana Zatz

## Abstract

Investigations on the concordance in monozygotic (MZ) as compared to dizygotic (DZ) twins may reveal if there is a genetic component increasing the susceptibility or resistance against an infectious disease. Here, we compared the concordance rates of SARS-CoV-2 infection in MZ versus DZ young twins who shared the same bedrooms and were equally exposed to the virus. The concordance rate was higher in the MZ group supporting a complex multifactorial inheritance responsible for SARS-Cov-2 infection.

---

The identification of genetic variants associated with higher susceptibility or resistance against SARS-Cov-2 infection has been the focus of innumerous studies around the world. Previous studies comparing monozygotic (MZ) and dizygotic (DZ) twins have been of paramount importance to identify if there is a genetic component driving the susceptibility to different infectious agents (1). If a condition depends mostly on the environment, it is expected that MZ and DZ twins will be equally affected. On the other hand, if the host genome plays a significant role, there is a higher concordance of outcomes in MZ when compared to DZ twins. By comparing MZ and DZ concordant and discordant twins, our group has shown that the occurrence of Zika Congenital Syndrome (ZCS) in discordant babies exposed to zika virus during pregnancy (one affected and one unaffected by ZCS clinical manifestations) was associated with host genetics (2). Such comparison is challenging for COVID-19, once the analysis requires a comparable exposure to the virus which is more difficult to control.

One of the major challenges in COVID-19 is also the inter-individual clinical variability ranging from complete absence of symptoms to severe and lethal outcomes even in younger individuals without comorbidities (3). In this context, since the spread of pandemic worldwide, reports of identical twins deceased due to COVID-19 within days apart drew the attention supporting the hypothesis of genetic components involvement in COVID-19. The first worldwide case of death from COVID-19, within three days apart in one pair of adult MZ was reported in April 2020 in the United Kingdom.

Here we report the outcome of COVID-19 in 10 pairs of young twins, 5 MZ and 5 DZ, living in Sao Paulo (the most populated Brazilian city). At least one sibling of each pair had confirmed COVID-19. These twins shared the same bedroom and were equally exposed to SARS-CoV-2 at home, without protection measures. In most cases, one or more relatives had confirmed COVID-19 with mild symptoms and transmitted the virus to the twins. The participants provided clinical details of the COVID-19 episode, including the laboratory tests and dates. Blood samples were collected, and we performed serological assays for SARS-CoV-2 IgA, IgG and IgM through enzyme-linked immunosorbent assay (ELISA) for the Receptor-Binding Domain (RBD) and Nucleocapsid (NP) proteins, at least four weeks after COVID-19 initial diagnosis.

Interestingly, four among the five MZ twin pairs were concordant for infection (both symptomatic or both asymptomatic) while among the five DZ twin pairs, four were discordant for infection (one infected and the other not) and just only one pair was concordant – based on clinical reports and confirmed by serology. **Table 1** shows the rates of concordance between MZ versus DZ twins, from our cohort.

**Table 1.**
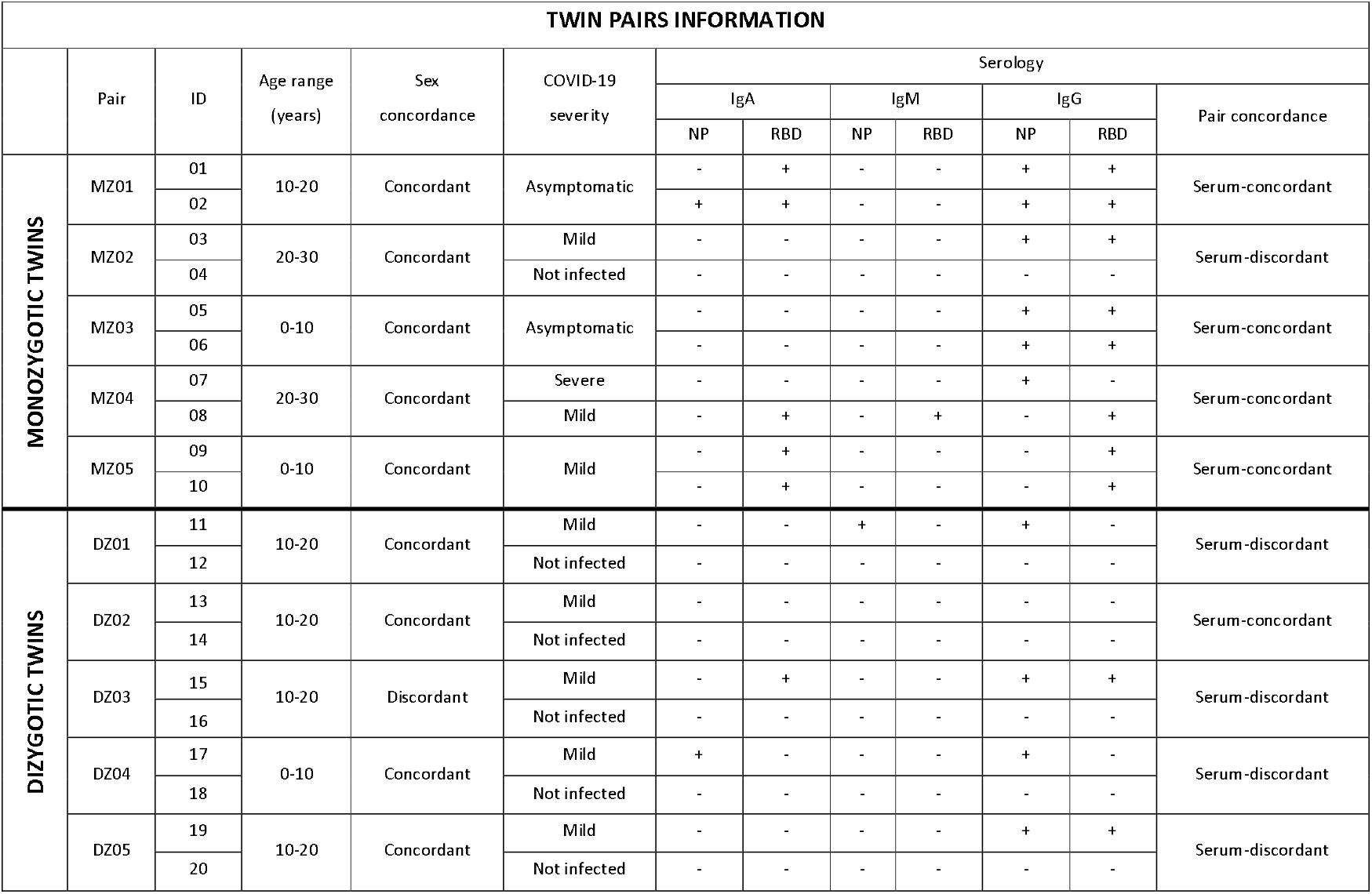
Demographical data, COVID-19 severity, and serological results of ELISA assay for SARS-CoV-2 IgA, IgG and IgM antibodies.

We also observed that in one pair of MZ twins, although both were infected and symptomatic, one was reinfected and displayed a severe illness while the other had an unique COVID-19 episode with mild symptoms (4). These observations are in accordance with a previous study reporting a case of COVID-19 in adult MZ twins who shared many of the same comorbidities, including type 2 diabetes mellitus, hypertension and overweight and had different disease courses and presentations (5). Curiously, the sibling with more complicated medical history experienced a milder and shorter COVID-19 course than the sibling who was hospitalized in ICU with a critical condition.

A case of simultaneous COVID-19 in adult MZ twins was also reported in the literature (6). In this one, the twins had similar health profiles without any chronic diseases or risk factors but had disparate clinical illnesses. One had a brief hospitalization while the other required intensive care and mechanical ventilation. In spite of being exposed to different environments, distinct outcomes were also reported for affected MZ twins even in congenital viral infections such as HIV (7) or cytomegalovirus (8), which supports a complex multifactorial inheritance.

In Brazil, at least seven cases of identical adult twins who died within days or weeks of differences were reported through the national media so far (**Table 2**). However, unlikely the first UK reported case, none of these Brazilian twins lived together or had any known health condition, reinforcing a genetic contribution behind SARS-CoV-2 susceptibility. Interestingly, 6 among the 7 pairs of twins are males, which is in accordance with the higher susceptibility and severity of COVID-19 in men than women (9, 10).

**Table 2.** Local reports of Brazilian monozygotic twins who died within days or weeks apart.

| Case | Sex | Age Range | Time Between Deaths (days) |
| --- | --- | --- | --- |
| 1 | M | 50-60 | 60 |
| 2 | M | 70-80 | 01 |
| 3 | M | 70-80 | 09 |
| 4 | M | 50-60 | 06 |
| 5 | M | 50-60 | 06 |
| 6 | M | 40-50 | 02 |
| 7 | F | 40-50 | 07 |

In conclusion, although these observations are derived from a small sample, the rate of discordance among DZ twins and the higher concordance in MZ gives further supports a complex multifactorial inheritance modulating the susceptibility or resistance against SARS-Cov-2 infection.

## Data Availability

Individual level data can be shared by authors upon reasonable request

## ACKNOWLEDGMENTS

The authors are extremely grateful for the participation and collaboration of the 20 twins and their families, the nurses for sample collection, the technical team, and the Fleury Laboratory for serology tests. Special thanks to Brazilian Senator Mara Gabrilli for financial support.

## ETHICS DECLARATION

This study was approved by the Committee for Ethics in Research of the Institute of Biosciences at the University of São Paulo (CAAE 34786620.2.0000.5464). The informed consent term was obtained from all participants.

